# Temporally Continuous Automated Sleep-Wake Classification Using Deep Learning

**DOI:** 10.64898/2025.12.03.25341129

**Authors:** Pranavan Somaskandhan, Henri Korkalainen, Timo Leppänen, Juha Toyras, Kerri Melehan, Warren Ruehland, Scott A. Sands, Dwayne L. Mann, Danielle L. Wilson, Philip I. Terrill

## Abstract

**Introduction:** Segmenting sleep into fixed 30-second epochs remains central to current sleep scoring practice, yet it imposes rigid boundaries that may not accurately reflect the true temporal sleep dynamics. We aimed to develop a deep learning-based, high-temporal-resolution sleep-wake classifier leveraging temporally continuous manual reference scoring without fixed epoch boundaries and transfer learning techniques to facilitate progress toward a more physiologically consistent sleep assessment.

**Methods:** Three independent datasets were utilized, of which two included sleep-wake scoring manually conducted in a temporally continuous manner. A U-Net based model was initially trained on a large dataset scored using 30-second epochs, with post hoc scoring modifications (*n*=2034). It was then fine-tuned via transfer learning using a subset of one of the datasets with temporally continuous scoring (*n*=39) and validated on both its holdout portion (*n*=40) and the other independent temporally continuous scoring dataset (*n*=20). Wakefulness and arousals were consolidated, acknowledging their shared physiological characteristics. Prediction confidence estimates were also generated.

**Results:** The model achieved overall concordance of 88.96% (*κ*=0.78) and 88.23% (*κ*=0.76) in the holdout and second independent evaluation dataset, respectively, with temporally continuous scoring. Correlation between 1-second automatic predictions and temporally continuous manual scoring was *r*=0.93 (*p*<0.001) for total sleep time and *r*=0.67 (*p*<0.001) for sleep-to-wake transition index.

**Conclusions:** These findings support the utility of our model in addressing key limitations of 30-second epoch-based scoring and progressing toward more physiologically consistent sleep-wake assessment by providing a practical basis for subsequent analyses. Misclassifications generally showed lower confidences, indicating additional value for targeted review.

**Statement of Significance:** Conventional sleep scoring remains constrained by fixed 30-second epochs, which may fail to capture the true temporal dynamics of the underlying changes between sleep and wakefulness. In this study, we used polysomnography data manually scored on a temporally continuous basis as the gold standard to develop and validate a deep learning model capable of classifying sleep and wakefulness-like states (consolidating wakefulness and arousal) at high temporal resolution without fixed 30-second epochs. The model demonstrated strong agreement with the gold standard, and as such, lays a practical foundation for deriving improved physiologically meaningful biomarkers of sleep fragmentation and continuity, with potential diagnostic and prognostic value and broad applicability toward a more precise and physiologically consistent sleep assessment.

## 1. Introduction

Sleep staging is an important step in diagnosing sleep disorders and in research investigating sleep physiology. However, conventional manual sleep staging remains laborious, time-consuming, and prone to human biases, leading to considerable inter-scorer variability [1–6]. To address these challenges, automated sleep staging systems have emerged as promising alternatives. In particular, modern deep learning-based approaches have achieved human-comparable performance [7–25]. While these systems have generally aimed to replicate and benchmark performance against trained humans scoring according to the American Academy of Sleep Medicine (AASM) guidelines [26], automation of sleep stage classification (particularly using deep-learning methods) presents the technical capability to move beyond certain constraints of conventional scoring practice [1,7,8,10].

A key constraint of current sleep scoring guidelines is the segmentation of sleep into fixed 30-second epochs—a convention first described by Loomis et al. in 1938 [27] and later standardized in 1968 based on paper-printed signals and data from healthy individuals [1,10,28]. While critical to the historic establishment of sleep research and clinical practice, this division does not capture true temporal dynamics of sleep-stage transitions, which rarely synchronize with 30-second epochs; and as such limits the ability to identify temporal associations with other important pathophysiological events. Furthermore, a 30-second epoch can often encompass features of both sleep and wakefulness, or two different sleep stages, yet must still be classified according to the predominant single stage in the epoch [26,29], likely contributing to inter-rater variability in sleep scoring [1,4,7,21,28]. In contrast, arousals events (as distinct from 30-second epoch scoring of wake) are scored on a temporally continuous basis when there is an abrupt shift in electroencephalography (EEG) frequency lasting at least 3 seconds, preceded by at least 10 seconds of stable sleep [26]. While arousals and wakefulness differ in duration, they share certain physiological features, such as increased EEG frequency and sympathetic nervous system activity reflecting elevated levels of cortical and subcortical activation [30,31]; and both may indicate pathological sleep disruption. Although guidelines suggest that qualifying arousals should be marked even within epochs scored as wakefulness [26,32], these overlapping characteristics may contribute to scoring dilemmas for human scorers, particularly during prolonged arousals, thereby contributing to poor inter-rater arousal scoring agreement [33,34]. In addition, this scoring distinction may introduce discrepancies in calculating clinical metrics such as the arousal index (ArI) [32].

Several studies have already made attempts to use automation of sleep scoring to move beyond the constraints of conventional sleep scoring. This includes classification at higher temporal resolution than 30-second epochs [12,22,24,35], and replacing categorical sleep staging with continuous sleep depth measures [36–40]. To date, these methods have either relied on manual reference annotations derived from conventional 30-second epoch-based scoring to train algorithms; and to the best of our knowledge, have not been directly validated against manual scoring annotated at a similar granularity as automated prediction.

This is likely due to the difficulty in reliably manually scoring sleep studies continuously in both time and across the full suite of five categorical sleep stages, or in both continuous time and continuous sleep depth. However, as a practical step toward a more physiologically consistent assessment of sleep, manual scoring in continuous time with a two-stage classification of sleep versus a combined wakefulness and arousal represents a more feasible approach.

As such, the aim of this study is to develop and systematically validate a deep learning-based high-temporal-resolution sleep-wake classification model that consolidates wakefulness and arousals. Uniquely, the gold standard for the training and validation of the algorithm utilizes polysomnography (PSG) data where sleep and “wakefulness-like” periods (encompassing both wakefulness state and arousals) were manually scored on a temporally continuous basis, without fixed epoch boundaries or durational limits. We hypothesize that a deep learning model based on the U-Net architecture, fine-tuned via transfer learning, would enable physiology-aligned, high-temporal-resolution sleep-wake classification demonstrating strong concordance with temporally continuous manual scoring. Ultimately, this study seeks to advance toward a more physiology-aligned approach to sleep scoring in both research and clinical applications.

## 2. Methods

The model training and validation included three datasets in total across initial training, fine-tuning, and evaluation. Two of these datasets, collectively encompassing 99 PSGs, comprised temporally continuous manual sleep-wake scoring. Given the limited availability of such data, we implemented a transfer learning approach. Specifically, we first pre-trained our deep learning model on a separate large PSG dataset labeled using conventional AASM arousal and sleep scoring guidelines, then fine-tuned it using a subset of one of our temporally continuous scoring datasets. The model was subsequently evaluated on the remaining holdout portion of that temporally continuous scoring dataset and on the other completely independent dataset with temporally continuous manual scoring.

### 2.1 Datasets

#### 2.1.1 Dataset Scored According to the AASM Guidelines - The Multi-Ethnic Study of Atherosclerosis (MESA)

The Multi-Ethnic Study of Atherosclerosis (MESA) Sleep, a large, publicly available dataset scored according to AASM guidelines, was utilized for initial model training [41,42]. MESA is a multi-center, longitudinal study investigating factors associated with cardiovascular disease in a diverse cohort of Black, White, Hispanic, and Chinese-American men and women. Participants aged 45–84 years were recruited during the baseline period between 2000–2002, with follow-ups. During the MESA Sleep Exam (2010–2012), 2237 participants underwent full overnight unattended PSG. For this study, we had access to 2060 PSGs. After excluding recordings with missing channels, no annotations, or incomplete annotations (i.e., those containing only wake or only wake and N2), the final dataset comprised 2034 recordings. From here on, this dataset is referred to as the MESA dataset.

#### 2.1.2 Datasets Scored Using Temporally Continuous Sleep-Wake Classification

The first of the two datasets scored using temporally continuous sleep-wake classification was acquired from Harvard, Boston. The original dataset comprised 96 PSGs from 85 participants, with 11 undergoing repeated PSGs from different visits (institutional ethics: 2023/HE000630). These full in-lab physiological studies primarily involved diagnosed obstructive sleep apnea (OSA; apnea–hypopnea index [AHI]≥5) patients (*n*=75), along with nine participants without OSA (AHI<5), and one participant who had an AHI<5 in the first visit but mild OSA (5≤AHI<15) in the second visit. PSGs included continuous positive airway pressure (CPAP) manipulation alongside periods of normal sleep. These segments were excluded from our analysis to avoid any bias introduced by CPAP manipulation. Recordings with both total sleep time (TST) below 120 minutes and total monitoring time under 240 minutes were excluded. Further exclusions included recordings with poor EEG quality or notable artifacts as determined through visual inspection, as well as those with missing channels of interest. The final dataset comprised 79 PSGs from 71 unique participants, with a mean (±standard deviation [SD]) AHI of 29.8 (±5.3). From here on, this dataset is referred to as the Harvard dataset.

Sleep stages in the Harvard dataset were initially scored according to the AASM guidelines [26]. In addition, a single expert scorer manually analyzed the PSGs and marked wakefulness-like periods (greater than 3 seconds duration) on a temporally continuous basis without an upper duration limit, using Spike software with a custom-built script to facilitate this scoring [43]. The scorer did not adhere to the epoch-based rule. This process ultimately yielded a temporally continuous manual sleep-wake classification for the dataset. It is noteworthy that the dataset did not include conventional arousal scoring as defined by AASM guidelines, as arousals were inherently captured through the temporally continuous scoring of wakefulness-like patterns.

The second dataset with temporally continuous sleep-wake scoring was acquired from the Sleep Laboratory at Austin Health, Victoria, Australia, and included 20 full in-lab PSGs conducted for suspected OSA (institutional ethics: 2024/HE000222). The data originated from an observational study that retrospectively reviewed diagnostic PSGs performed between March and May 2019 [32]. The dataset included four participants without OSA (AHI<5) and 16 with AHI≥5. The mean (±SD) AHI for these studies was 21.9 (±22.5). From here on, this dataset is referred to as the Austin dataset.

The Austin dataset was initially scored manually according to the AASM guidelines, including both sleep stage and arousal annotations. In addition, temporally continuous sleep-wake scoring was performed [32]. This refined scoring was conducted by an experienced sleep scientist, who manually reanalyzed the PSGs and marked the EEG as either sleep or wakefulness-like on a temporally continuous basis. The scorer added an additional channel adjacent to the existing signals post-recording and used an event marker to manually annotate periods of sleep without adhering to any fixed epochs. The identification of sleep onset was based on the attenuation and loss of alpha rhythm, the emergence of theta activity, and the presence of slow rolling eye movements in the electrooculography (EOG).

Figure 1 presents a 2-minute PSG example (four consecutive 30-second epochs), highlighting how temporally continuous manual scoring of sleep and wakefulness-like periods appears compared to conventional AASM-based sleep-wake scoring. It illustrates that temporally continuous scoring captures all sleep-to-wake and wake-to-sleep transitions reflected in PSG signals with greater temporal precision, whereas the AASM-based approach, by definition, assigns a single representative state to each 30-second epoch.

**Figure 1:**
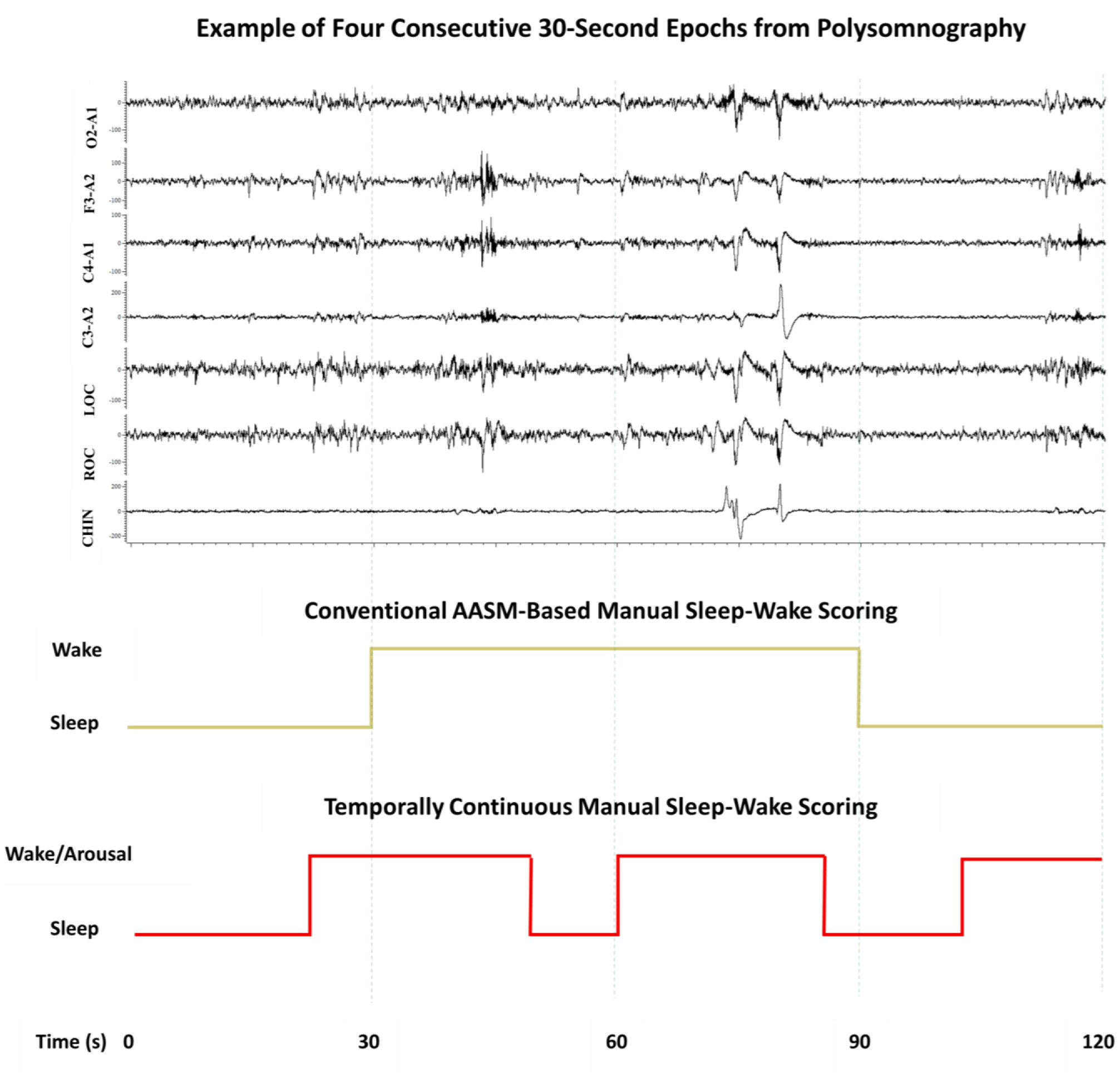
Illustration of how temporally continuous manual scoring of sleep and wakefulness-like periods differs from conventional AASM-based sleep-wake scoring. The top panel shows four consecutive 30-second epochs of PSG signals, including EEG (O2–A1, F3–A2, C4–A1, C3–A2), EOG (LOC, ROC), and chin EMG (CHIN). The middle panel displays the corresponding AASM-based manual sleep-wake scoring, where each 30-second epoch is assigned a single stage based on predominant activity. According to this conventional approach, the first epoch is scored as sleep, followed by two epochs marked as wake, and the final epoch as sleep. In contrast, the bottom panel presents temporally continuous manual scoring, which marks all transitions between sleep and wakefulness-like states with temporal precision, without being constrained by fixed epoch boundaries or durational thresholds. PSG = polysomnography, AASM = American Academy of Sleep Medicine, EMG = electromyography, EOG = electrooculography, EEG = electroencephalography. Vertical grey lines represent epoch boundaries.

### 2.2 Neural Network Architecture

We utilized a modified version of the U-Sleep architecture [12], which is a fully convolutional, feed-forward neural network designed for sleep staging. The rationale for selecting this architecture was its previously demonstrated generalizability across multiple datasets [12]. Moreover, U-sleep is built based on the U-Net architecture [44], which is inherently suited for high-temporal-resolution predictions [44,45]. The base architecture was simplified and fully retrained using our data, also incorporating shorter epoch durations. Similar to the original U-Sleep study, classifications were performed using input signals from any available pairs of common EEG and EOG electrodes regardless of sampling rate, which were internally resampled to 128 Hz. The model was designed to output both sleep-wake scoring and corresponding prediction probabilities, indicating the model’s estimated likelihood of each state. These probabilities reflect the model’s confidence in assigning each time point to either sleep or wakefulness-like and are derived from the softmax output.

Supplementary Figure S1 presents an overview of the architecture, with further technical details, including important modifications, provided in the supplementary document under the section "Details of Neural Network Architecture".

### 2.3 Initial Model Training and Evaluation Using MESA

The MESA dataset (*n*=2034) was divided into training (*n*=1524; 75%), validation (*n*=204; 10%), and test (*n*=306; 15%) sets. Sleep stages scored according to the AASM guidelines using 30-second epochs were first reclassified as sleep (for epochs labeled N1, N2, N3, or REM) or wake. Initial model training and evaluation were then conducted using two approaches:

1. **Conventional method:** Binary sleep-wake classification directly based on conventional AASM guidelines.
2. **With scoring modifications:** Classification of sleep and wakefulness-like patterns using heuristic post hoc scoring modifications to integrate arousals scored according to the AASM guidelines.

#### 2.3.1 MESA Scoring Modifications

Considering the inherent differences between fixed 30-second epoch-based and temporally continuous scoring, two post hoc adjustments were made to the MESA manual sleep annotations to align them more closely with the ultimately intended temporally continuous sleep-wake scoring. These involved incorporating arousals into the sleep-wake hypnogram and applying a heuristic correction where needed to adjust the sleep-to-wake transition points. While these modifications were not intended to fully replicate our gold standard temporally continuous scoring, they provide a closer approximation and serve as a foundation for transfer learning.

##### Arousal Incorporation

Independent of duration, arousal events were treated like wake, acknowledging their physiological similarities, and were overlaid onto the sleep-wake hypnogram. Specifically, sleep segments overlapping with arousals were converted to wakefulness-like at a 1-second resolution. If an arousal was marked within a 30-second epoch labeled as wakefulness according to AASM rules, then the corresponding wake segment was left unchanged. Figure 2A provides an illustrative example of this modification.

**Figure 2:**
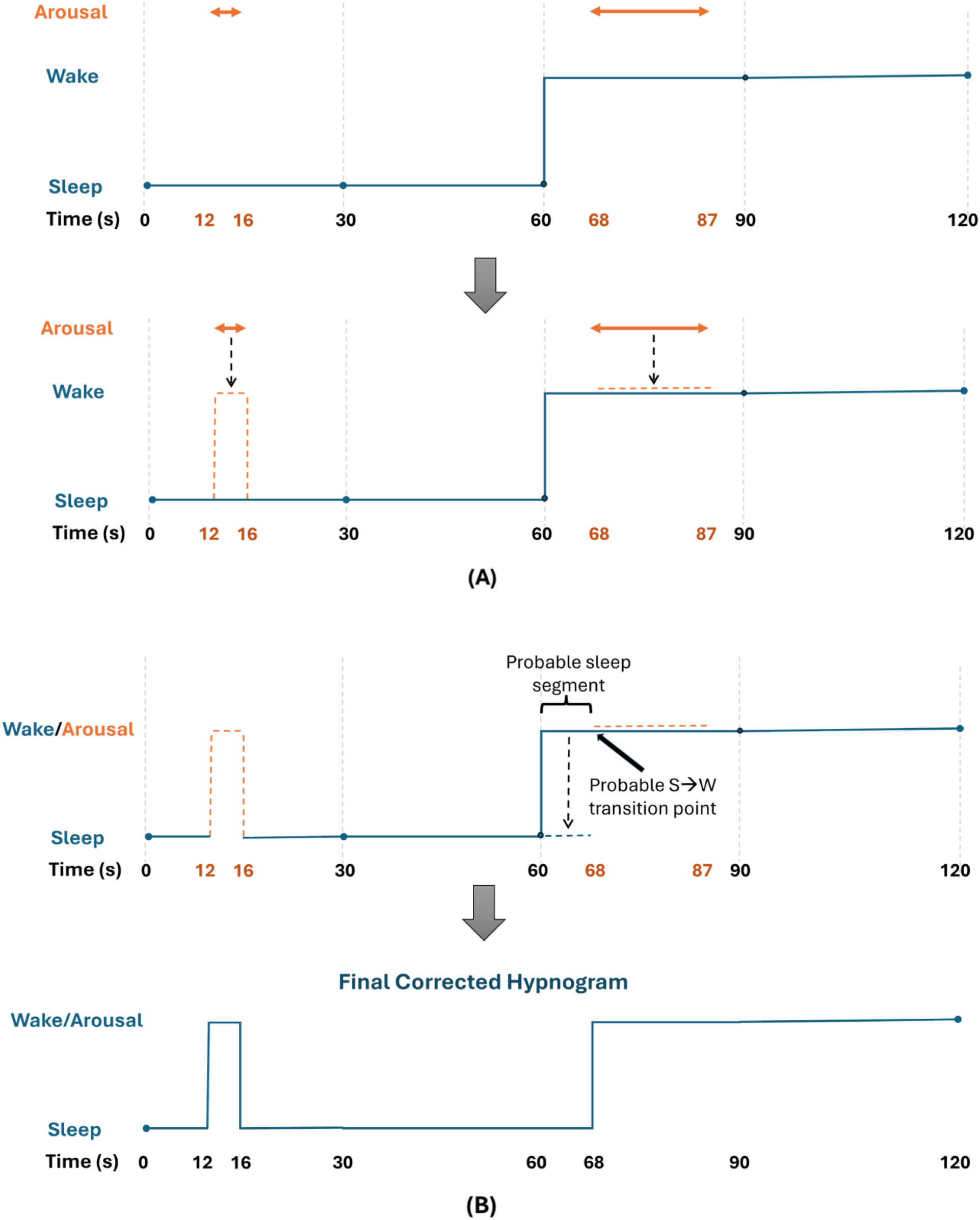
Illustrative examples of the Multi-Ethnic Study of Atherosclerosis (MESA) dataset post hoc scoring modifications. **(A)** Arousal incorporation step: Top panel shows a 2-minute sleep-wake hypnogram along with separately marked arousals, according to AASM guidelines. Second panel shows how these arousals are overlaid onto the sleep-wake hypnogram and incorporated. When an arousal occurs during a marked sleep epoch, the corresponding segment is converted to wakefulness-like. Arousals marked in wake epochs do not alter the hypnogram per se, however these arousals are used in the heuristic correction. **(B)** Heuristic correction step: The arousal marked between 68-87 seconds appears in the first half of that wake epoch, immediately following a sleep epoch. It is likely that the actual S→W transition occurred at the onset of arousal rather than at preceding epoch boundary. Thus, the portion of wake preceding the arousal within that epoch is heuristically corrected as sleep. Bottom panel shows the final corrected hypnogram. AASM = American Academy of Sleep Medicine, S→W: sleep-to-wake.

##### Heuristic Correction

Fixed 30-second epoch-based scoring may not always align with actual sleep transitions, leading to potential misalignments in transition localization. To address some of these sleep-to-wake transition misalignments and better approximate the temporally continuous scoring, an additional step was applied after arousal incorporation to refine transition timing where necessary. Specifically, when an arousal occurs within the first half of a wake epoch that immediately follows a sleep epoch, it is likely that the actual sleep-to-wake transition occurred at the onset of the arousal rather than at the preceding 30-second epoch boundary. Since conventional scoring assigns wakefulness to an entire epoch if wake features predominate, this may overestimate wake duration and misrepresent such transition points.

To account for this, a heuristic correction step was applied under the following three conditions:

1. The arousal was marked in an epoch that was manually scored as wakefulness.
2. The arousal occurred in the first half of that wake epoch.
3. The preceding epoch was scored as sleep.

If all three criteria were met, the portion of wake preceding the arousal within that 30-second epoch was heuristically corrected and reassigned as sleep (Figure 2B).

Models were systematically evaluated across multiple epoch durations: 1-, 3-, 5-, 10-, 15-, and 30-second. For this study, we focus on the shortest 1-second resolution with the modified scoring approach. Input signals from MESA comprised Fz-Cz, Cz-Oz, C4-M1, E1-FPz, and E2-FPz channels. These signals included internal high-pass and low-pass filters at 0.3 Hz and 35 Hz, respectively, and were resampled from 256 Hz to 128 Hz using polyphase filtering. Segments outside the manually scored periods at the beginning and end of recordings were trimmed during training. To handle noisy values, we relied on the outlier-robust clipping mechanism from the original U-Sleep study [12].

### 2.4 Transfer Learning

To adapt and further fine-tune the model for temporally continuous scoring, transfer learning was employed. This artificial intelligence technique involves pre-training a model on a large, related dataset and subsequently fine-tuning it using limited data for the target task, helping maximize the utility of our uniquely scored data. The MESA pre-trained 1-second model with arousal incorporation and heuristic correction steps served as the base model. The Harvard dataset was divided into two approximately equal subsets: 39 PSGs for transfer learning (further fine-tuning) and 40 PSGs for final evaluation. To avoid biases, PSGs from the same participant were exclusively assigned to the fine-tuning subset (8 participants had repeat studies). A four-fold cross-validation approach was used during transfer learning, with three folds for training and one for validation. The final model was then evaluated on the Harvard holdout set (*n*=40). Additionally, the model’s generalizability was further assessed on the external Austin dataset (*n*=20). Further technical details of the transfer learning approach are provided in the supplementary document under the section "Details of Transfer Learning Procedure".

### 2.5 Model Evaluation

Our primary metrics for evaluating model performance were percentage concordance, Cohen’s kappa (*κ*) [46], sensitivity, and specificity. These were computed for the Harvard holdout and the Austin Datasets; and for comparison, reported for the Harvard fine-tuning subset. We also applied the MESA-trained model directly on our evaluation data (i.e., Harvard holdout and Austin datasets) before fine-tuning to determine if the transfer learning improves the model performance. Furthermore, we compared the TST and the sleep-to-wake transition index (i.e., the total number of sleep-to-wake transitions per hour) using correlation plots and Bland-Altman analysis.

We also conducted a number of further analyses that provide additional insights into technical performance and clinical utility. Firstly, to assess whether the misclassifications were predominantly confined to shorter durations or extended across longer segments, we generated histograms of the duration of continuous misclassified segments, separately for both the Harvard holdout and Austin datasets. Secondly, in addition to a prediction of state (sleep or wakefulness-like), the deep learning algorithm was configured to provide a prediction probability. The mean prediction confidence was calculated for each participant by averaging the model’s prediction probabilities across the night; and to examine the relationship between prediction certainty and classification performance, correlations between the mean prediction confidence and performance metrics were analyzed. Histograms of the prediction confidences for misclassified 1-second segments (expressed as a percentage of total segments within each bin) were generated in both evaluation datasets to provide insights into confidence levels associated with misclassification.

## 3. Results

### 3.1 Demographic Statistics of Datasets

Table 1 presents a summary of key demographics and characteristics of datasets.

**Table 1:** Demographics and characteristics of datasets.

| Dataset |  | MESA | Harvard | Austin |
| --- | --- | --- | --- | --- |
| Records |  | 2034 | 79 | 20 |
| Participants |  | 2034 | 71 | 20 |
| Scoring protocol |  | Conventional only | Conventional <sup>#</sup> + Temporally Cont. <sup>*</sup> | Conventional + Temporally Cont. <sup>*</sup> |
| Age (Years) [Mean ± SD] | | 68.5 ± 9.2 | 52.5 ± 13.0 | 58.9 ± 14.1 |
| BMI (kg/m<sup>2</sup>) [Mean ± SD] | | 28.6 ± 5.5 | 32.1 ± 6.5 | 33.3 ± 6.8 |
| Sex % (Female / Male) |  | 53.8 / 46.2 | 32.4 / 67.6 | 40.0 / 60.0 |
| TST (Minutes) [Mean ± SD] | Conventional | 360.7 ± 81.9 | 219.7 ± 93.8 | 225.4 ± 109.7 |
| | Temporally Cont. | - | 204.1 ± 89.0 | 246.0 ± 106.3 |
| Sleep Efficiency (%) [Mean ± SD] | Conventional | 79.7 ± 12.7 | 63.4 ± 19.6 | 50.2 ± 23.3 |
| | Temporally Cont. | - | 58.7 ± 18.4 | 55.2 ± 24.0 |
| AHI (Events/Hour) [Mean ± SD] | Conventional | 23.4 ± 19.0 | 30.6 ± 25.4 | 21.9 ± 22.5 |
| | Temporally Cont. | - | 33.8 ± 29.6 | 19.9 ± 20.3 |

SD = standard deviation, BMI = body mass index, TST = total sleep time, AHI = apnea-hypopnea index, Temporally Cont. = temporally continuous.
<sup>#</sup> The Harvard dataset included only conventional sleep stage scoring and did not provide arousal scoring according to AASM guidelines, as arousals were inherently captured through the temporally continuous scoring of wakefulness-like patterns.
<sup>\*</sup> A temporally continuous scoring protocol in this study refers to manual scoring of sleep and wakefulness-like (encompassing both wakefulness state and arousals as a single category) periods that was conducted in a temporally continuous manner without fixed epoch boundaries or durational limits.

### 3.2 Initial Model Training and Evaluation in the MESA Dataset

During preliminary training on the MESA dataset with arousal incorporation and heuristic correction steps, the automatic 1-second sleep-wake classification model achieved concordances of 88.62% (*κ*=0.79) and 90.89% (*κ*=0.81) with modified reference scoring in the training and validation sets, respectively. In the MESA holdout set, it reached a concordance of 91.04% (*κ*=0.82), with sensitivity and specificity values of 88.85% and 92.90%, respectively, for identifying wakefulness-like patterns. When directly evaluated on datasets with temporally continuous manual scoring, the MESA-trained model achieved concordances of 86.21% (*κ*=0.72) and 86.66% (*κ*=0.74) in the Harvard holdout (*n*=40) and the Austin (*n*=20) datasets, respectively.

### 3.3 Post-Transfer Learning Evaluation in Datasets with Temporally Continuous Sleep-Wake Classification

After transfer learning using the Harvard data subset (*n*=39), the refined model demonstrated improved concordance with temporally continuous manual scoring. In the unseen Harvard holdout set, the model achieved an overall concordance of 88.96% (*κ*=0.78), with a sensitivity of 85.90% and a specificity of 91.51%. Similarly, in the Austin data, the model achieved an overall concordance of 88.23% (*κ*=0.76), with a sensitivity of 89.95% and a specificity of 86.71%.

Figure 3A illustrates the distribution of percentage agreement in the Harvard holdout and Austin datasets after transfer learning, while Figure 3B presents the corresponding distributions of Cohen’s kappa agreement. The boxplots demonstrate that the model performs similarly in both datasets, as reflected by similar medians and interquartile ranges.

**Figure 3:**
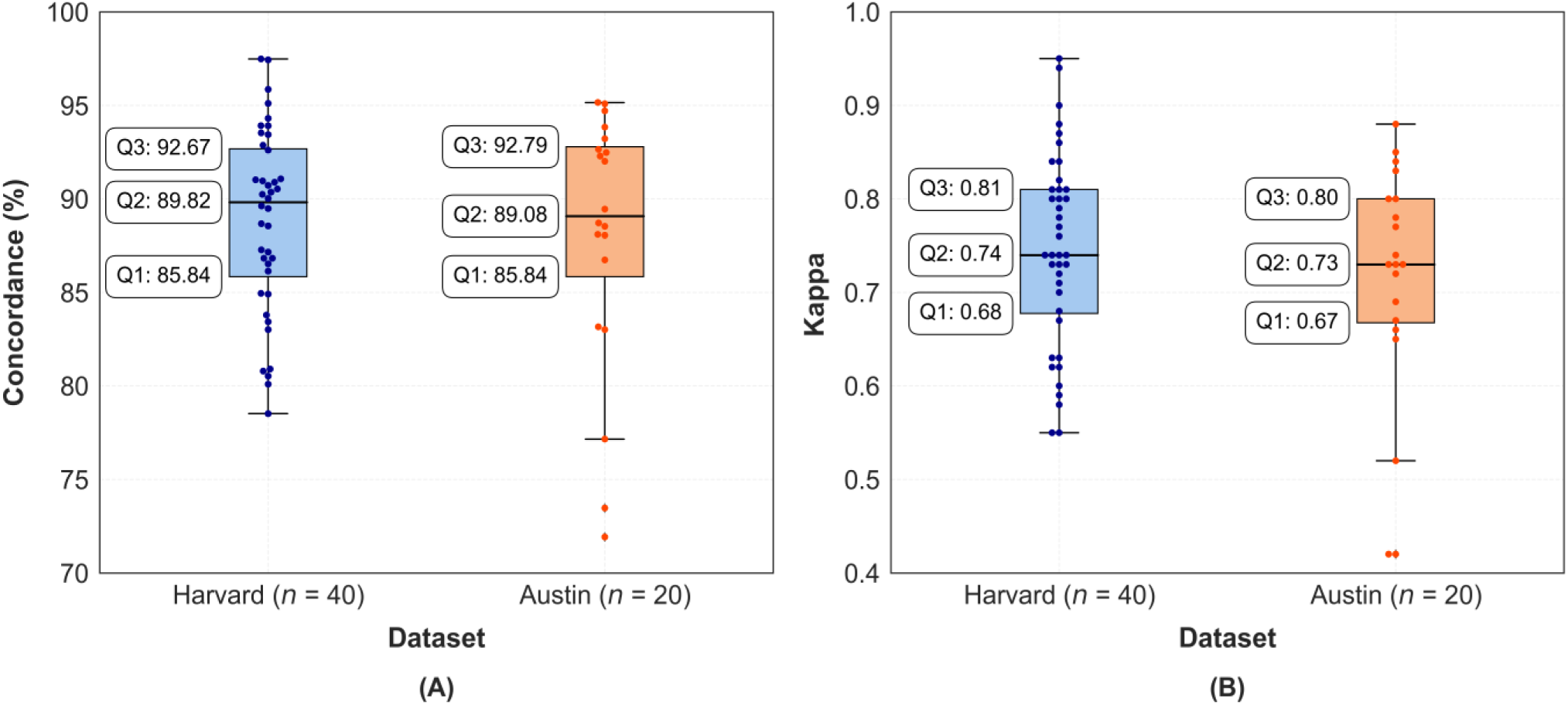
Participant-wise performance distributions in the Harvard holdout and Austin datasets after transfer learning: (A) Percentage agreement and (B) Cohen’s kappa agreement. Q1: First quartile, Q2: Median, Q3: Third quartile.

We further examined how TST, determined by the automatic method, compared with temporally continuous scoring and found a strong correlation of 0.93 (*p*<0.001) between the two methods (Figures 4A and 4C). Similarly, we compared sleep-to-wake transition indices and observed a correlation of 0.67 (*p*<0.001; Figures 4B and 4D). Additionally, given the minimum duration criteria for manual arousal scoring in the Harvard dataset, we applied a post-hoc heuristic correction to 1-second standalone sleep or wake predictions (i.e., a 1-second segment of one class surrounded by the opposite class). These segments were smoothed if the mean prediction probability for that class, calculated over a 3-second window centered on the segment, was below 0.5. Otherwise, the entire 3-second window was reassigned to the class of that 1-second segment. This correction improved the correlation between the methods for the sleep-to-wake transition index to 0.72 (*p*<0.001) and reduced the mean difference to 0.19 in the Bland-Altman analysis (see Supplementary Figure S2). All other results remained largely unchanged.

**Figure 4:**
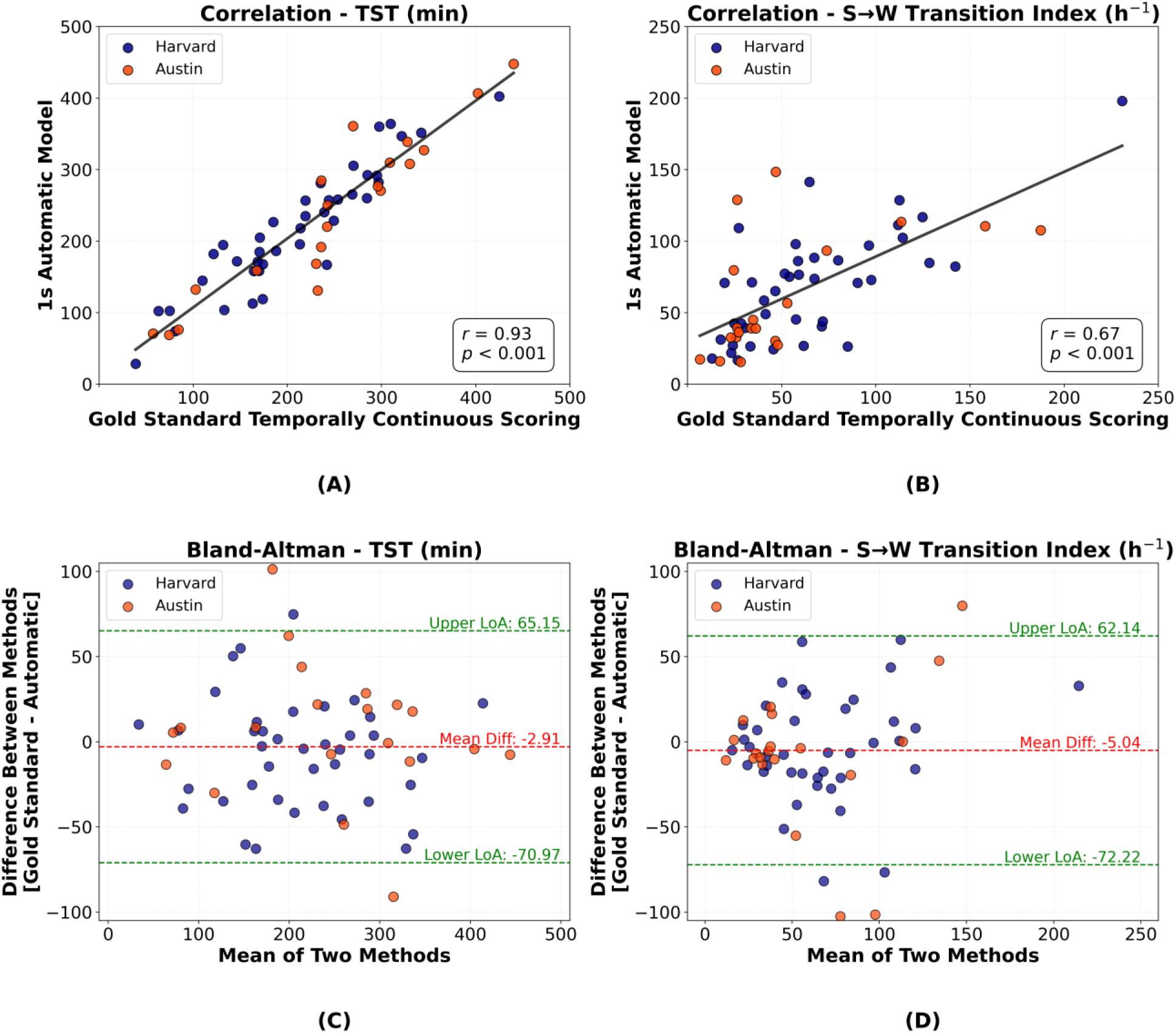
Analyses of TST and sleep-to-wake transition indices between the 1-second automatic method and the gold standard temporally continuous scoring. (A) Correlation plot of TST (r=0.93, p<0.001). (B) Correlation plot of sleep-to-wake transition index (r=0.67, p<0.001). (C) Bland-Altman plot for TST comparison. (D) Bland-Altman plot for sleep-to-wake transition index. TST = total sleep time, S→W = sleep-to-wake.

A 15-minute sleep-wake hypnogram example from a participant in the Harvard holdout set is presented in Figure 5A to visually compare scoring. It includes (I) a hypnogram based on manual AASM annotations where arousals were incorporated into sleep-wake scoring, (II) a hypnogram based on temporally continuous manual scoring, (III) a hypnogram predicted by the automatic model, and (IV) sleep-wake probability distributions corresponding to the predicted hypnogram. Figure 5B illustrates a similar example from the Austin dataset. In both cases, we observe that the model predictions closely align with temporally continuous scoring.

**Figure 5:**
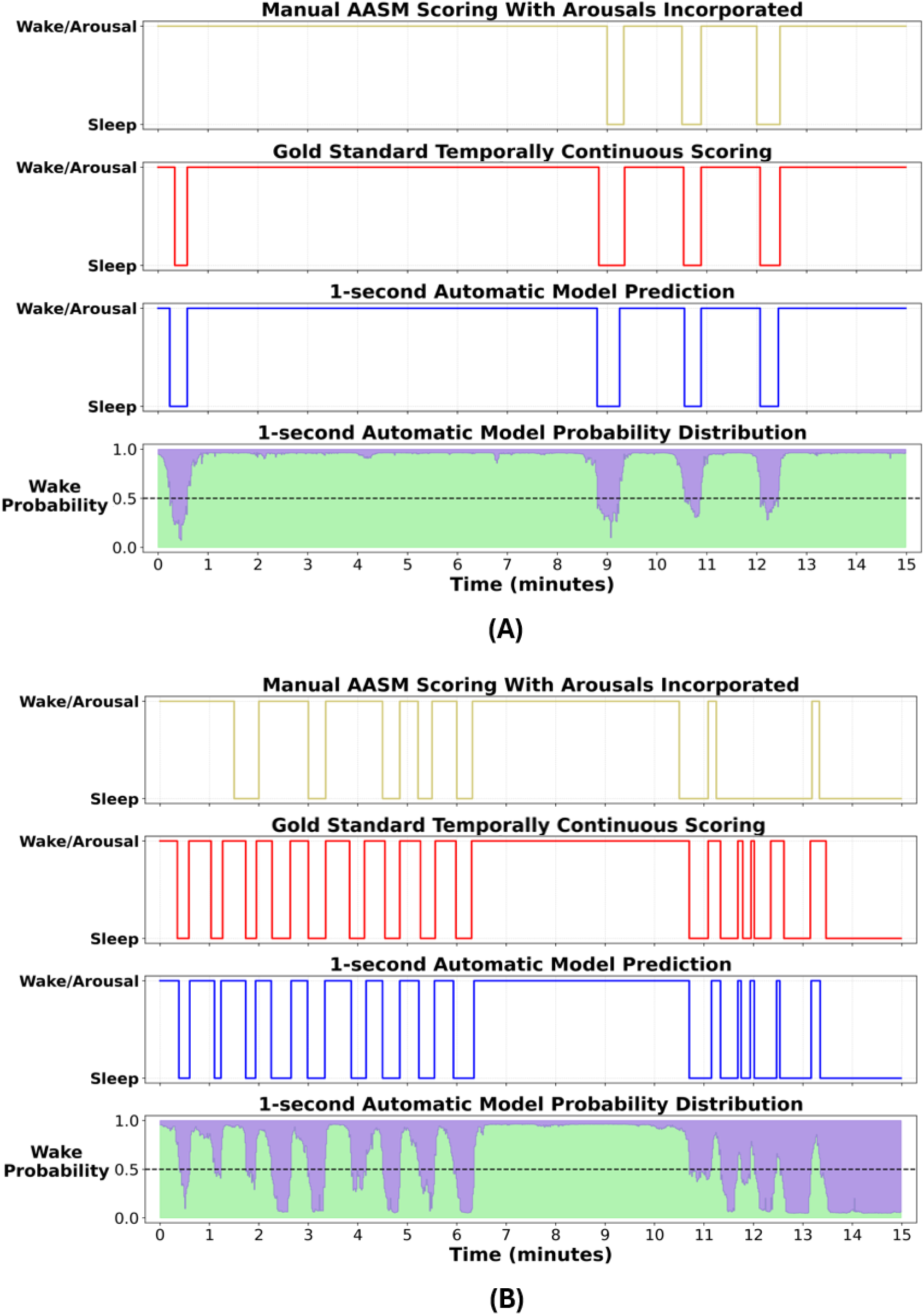
15-minute sleep-wake hypnogram examples from (A) a participant from the Harvard holdout set and (B) a participant from the Austin dataset. (I) hypnogram based on manual American Academy of Sleep Medicine (AASM) annotations where arousals were incorporated into sleep-wake scoring, (II) hypnogram based on our gold standard temporally continuous scoring, (III) hypnogram predicted by our 1-second automatic model, and (IV) sleep-wake probability distributions corresponding to our predicted hypnogram (green color coding denotes the probability of wake for each time point, while purple indicates the inverse sleep probability).

The histograms of the duration of misclassified segments illustrated that most misclassifications were concentrated in shorter durations (Figure 6).

**Figure 6:**
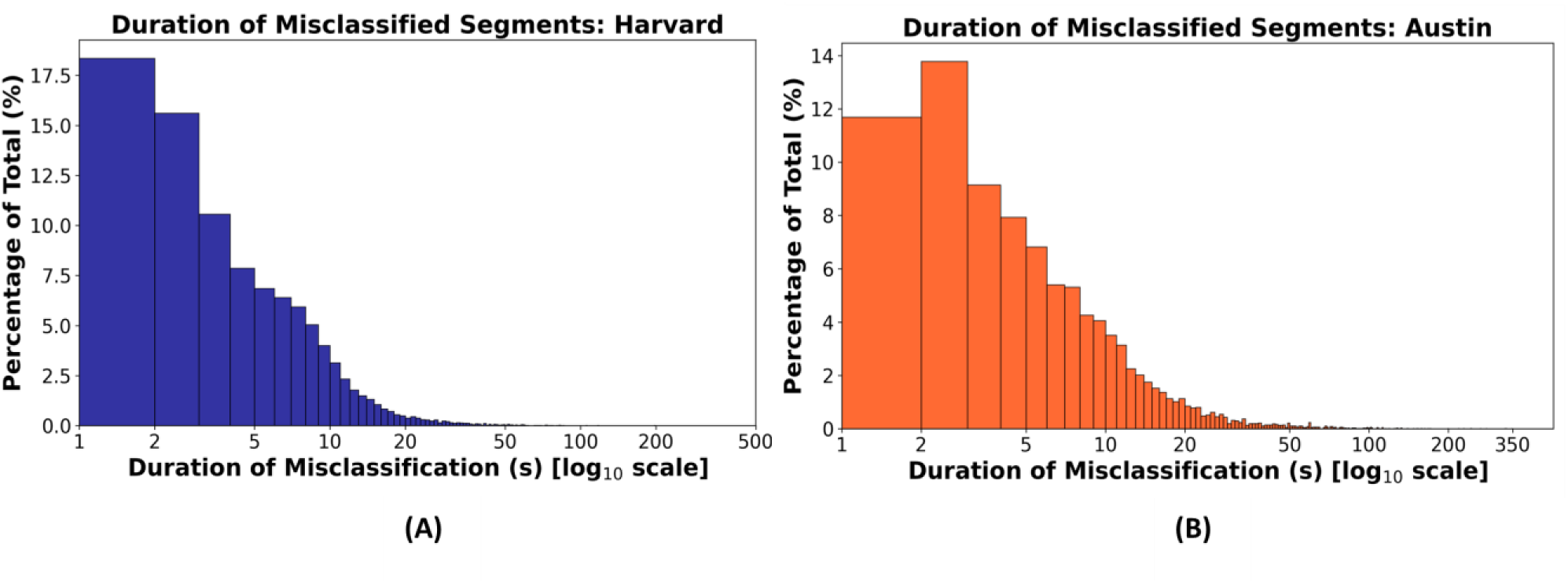
Histograms of the duration of misclassified segments in the Harvard holdout and Austin datasets. Histograms show the percentage distribution of misclassified segments (defined as discrepancies between the automatic predictions and temporally continuous manual scoring), grouped by duration, separately for the **(A)** Harvard holdout and **(B)** Austin datasets. Each bar indicates the proportion of total errors corresponding to a specific duration (in seconds). These plots demonstrate that most misclassifications were short in duration.

A moderate correlation of 0.63 (*p*<0.001; Figure 7A) was observed between mean prediction confidence and percentage concordance, while a similar correlation of 0.58 (*p*<0.001; Figure 7B) was found with kappa. These results suggest that generally the model tends to be more confident in its predictions when it achieves higher agreement with the manual scoring. Further, we investigated prediction confidences for misclassified 1-second segments. Figure 8 presents histograms of the prediction confidences for misclassified segments (expressed as a percentage of total segments within each bin) in both datasets separately. The histograms indicate that most misclassifications occurred with lower confidence levels, which suggests that model’s predictions were less certain during most discrepancies, reflecting better model behavior.

**Figure 7:**
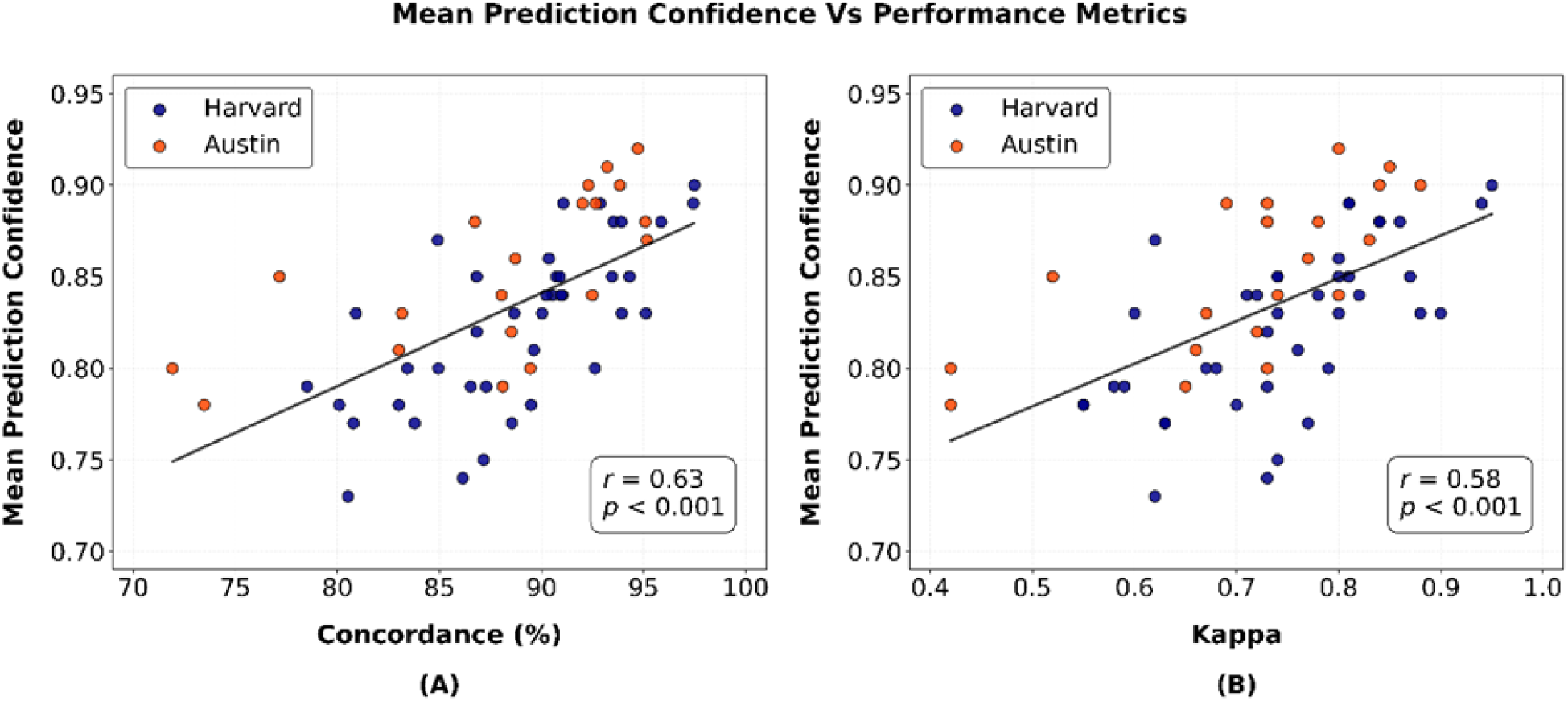
Scatter plots showing the relationship between mean prediction confidence and performance metrics: (A) percentage agreement (r=0.63, p<0.001) and (B) Cohen’s kappa (r=0.58, p<0.001) across the Harvard holdout and Austin datasets. The regression lines represent the trends in both datasets.

**Figure 8:**
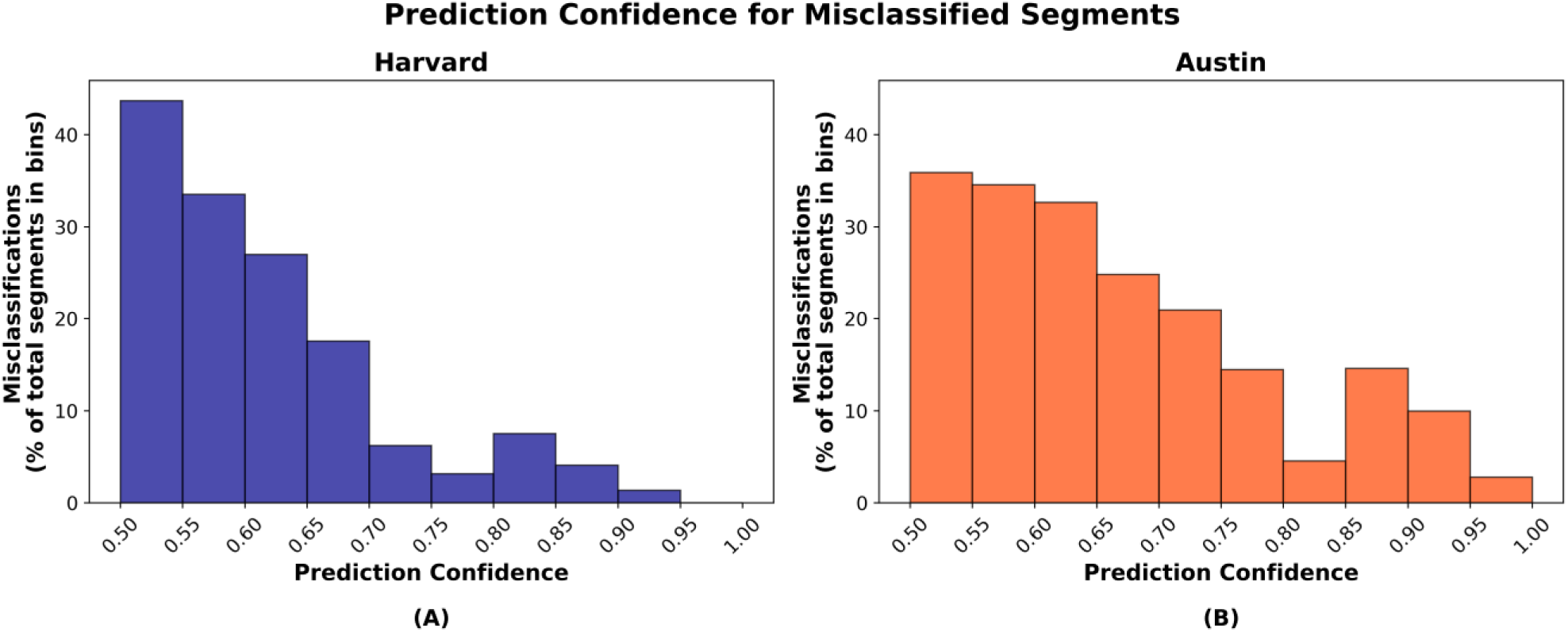
Histograms of the prediction confidences for misclassified 1-second segments, expressed as a percentage of total 1-second segments within each confidence bin. (A) presents results from the Harvard holdout dataset and (B) presents results from the Austin dataset.

## 4. Discussion

The objective of this study was to develop and validate a deep learning-based high-temporal-resolution sleep-wake classification model that also consolidates wakefulness and arousals into a “wakefulness-like” classification. We hypothesized that a U-Net based architecture, fine-tuned via transfer learning, would enable physiology-aligned, high-temporal-resolution sleep-wake classification, demonstrating strong concordance with temporally continuous manual scoring. Our findings substantiated this hypothesis, as the model achieved high concordance with our gold standard temporally continuous scoring (i.e., manual sleep-wake scoring without epoch boundaries) not only in the Harvard holdout set (88.96%, *κ*=0.78) but also in the completely independent Austin dataset (88.23%, *κ*=0.76). Participant-wise performance distributions were similar across both datasets, with consistently high values (Figure 3).

Our work builds upon and addresses key gaps in existing approaches aimed at moving beyond the constraints of conventional 30-second epoch-based sleep scoring. Younes et al. [36,37] proposed a novel method to evaluate sleep on a continuous scale using the odds ratio product (ORP) as a measure of sleep depth. This method has demonstrated value in capturing sleep-related physiological variation, as well as associations with sleep disorders such as OSA, insomnia, excessive sleepiness, and perceived poor sleep quality [36,37]. However, despite its promise, this approach has not been directly validated against alternative manual annotations scored in a comparable manner. Such validation remains extremely challenging due to the difficulty in manually scoring sleep continuously in both time and depth. Practical barriers may also exist, such as the need for widespread standardization and integration into clinical workflows, which might involve considerable cost, adaptation, and interpretability-related challenges [10]. Other non-categorical scoring systems [38–40] face similar limitations, highlighting the need for solutions that overcome current shortcomings while potentially allowing easier clinical adaptation.

High-frequency sleep scoring systems leveraging deep learning have demonstrated potential for improved temporal precision [12,22,24,35]. The U-Sleep architecture by Perslev et al. [12,35] is inherently capable of scoring sleep at high temporal resolutions. However, it lacks validation against true physiology-aligned scoring to thoroughly assess the reliability of high-frequency sleep staging. Other finer-resolution scoring systems share similar constraints. Machine learning-based clustering approaches have also been explored to reframe sleep staging [47,48], although their primary aim was not to eliminate fixed epoch boundaries. In contrast to these works, our study developed a physiology-aligned, high-temporal-frequency sleep-wake classification approach that was systematically validated using datasets manually scored on a temporally continuous basis without epoch constraints. We acknowledge that this was practically achievable by reducing the number of classes from five in conventional AASM scoring (N1, N2, N3, REM, wake) to two (sleep and consolidated wake/arousal).

Direct performance benchmarking of our model is limited due to the absence of similar comparable methods. However, we compared our results with existing literature on AASM-based binary sleep-wake and arousal classifications separately. While our model’s agreement appears slightly lower than some prior binary sleep-wake classifiers based on EEG [18,49,50], this is expected given the broader scope of our task. Unlike these, our model consolidates wakefulness and arousals and still achieves strong, comparable performance. When considering conventional arousal scoring, its inter-rater reliability is notoriously poor in both manual and automated scoring, with kappa values typically around just 0.40 [33,34,51–53]. Conversely, our model demonstrated high concordance with temporally continuous manual scoring in what may represent a more complex task, accurately capturing both short- and long-duration wakefulness-like patterns. Some automated studies have also attempted simultaneous sleep staging and arousal detection [54,55], but these remain constrained by adherence to conventional epoch-based rules and reliance on training data with potentially suboptimal arousal label quality.

Beyond concordance, additional metrics helped characterize model robustness. Specifically, strong correlations were observed between model’s predictions and temporally continuous manual annotations for key sleep metrics, such as TST (*r*=0.93, *p*<0.001) [Figures 4A and 4C] and sleep-to-wake transition index (*r*=0.67, *p*<0.001) [Figures 4B and 4D]. The histograms of the duration of misclassified segments further demonstrated that most misclassifications were confined to shorter durations, confirming that the automatic model rarely deviated from the temporally continuous sleep-wake scoring for extended periods (Figure 6). Collectively, these findings validate the algorithm’s capability to reliably capture the underlying sleep-wake temporal dynamics.

### 4.1 Physiological Insights and Clinical Applications

The developed algorithm offers valuable physiological insights and potential clinical applications. The algorithm closely aligned with the temporal continuity of sleep-wake transitions; and showed well-balanced sensitivity and specificity, underscoring its potential to accurately detect both short and long sleep and wake-like patterns. Despite not classifying the full suite of AASM sleep states, this algorithm is particularly well-suited for assessing sleep disruption and fragmentation, which is a hallmark of conditions such as sleep-disordered breathing. In particular, the high temporal resolution and consolidation of arousals and wakefulness allow the temporal association with other clinically important events (such as apnea or hypopnea) to be investigated. Additionally, specific use cases such as the Maintenance of Wakefulness Test (MWT) and the Multiple Sleep Latency Test (MSLT) [56], could significantly benefit from the algorithm’s ability to detect microsleeps and brief intrusions of sleep prior to scored sleep onset. This is particularly relevant in these tests, where conventional scoring focuses on sleep latency to the first scored 30-second sleep epoch, potentially overlooking early, subtle vigilance lapses that may better capture early episodes of impaired wakefulness or daytime sleepiness.

The ability to estimate TST more accurately, count discrete sleep-to-wake transitions, and consolidate wakefulness and arousals without durational limits opens new opportunities to derive physiologically meaningful biomarkers that may be useful in predicting a range of clinical outcomes. One clear example is the sleep-to-wake transition index derived from our algorithm, which precisely quantifies the number of unique times that a person shifts from sleep to wake. Neither conventional 30-second epoch-based hypnograms nor the arousal index captures this directly, and simply combining them would risk double-counting or missing transitions when wake epochs and arousal events overlap. In contrast, this algorithm allows for a more accurate measure of sleep fragmentation by directly identifying each shift from sleep to wakefulness-like states. Such physiologically consistent biomarkers, in turn, may have high value in predicting clinical outcomes such as vigilance, excessive daytime sleepiness, cardiovascular disease, or all-cause mortality. While evaluating these associations is beyond the scope of the present study, our method provides a practical foundation for future work that may potentially lead to improved diagnostic and prognostic value for certain clinical conditions.

### 4.2 Technical Insights

Our work provides valuable technical contributions toward refining and scaling automated sleep scoring methods, particularly through the utilization of transfer learning. Performing temporally continuous manual sleep-wake scoring at a large scale remains challenging and time-consuming. The transfer learning approach enabled our model to adapt to temporally continuous manual scoring by leveraging only a relatively small amount of such data for fine-tuning. Our work further highlights the promise of transfer learning in sleep research, allowing algorithms to retain knowledge from prior training and adapt through fine-tuning to even smaller datasets with varying scoring criteria, demographic characteristics, or clinical conditions, while reducing extensive retraining. Furthermore, through the integration of prediction confidences, the algorithm offers a practical means to assess classification certainty and guide targeted manual review. Our findings demonstrated that mean prediction confidences were moderately correlated with performance metrics (Figure 7), and misclassifications generally showed lower prediction confidences, reflecting better model behavior (Figure 8). These low-confidence misclassifications may serve as simple flags for further inspection.

### 4.3 Methodological Limitations and Future Directions

While this study advances beyond 30-second epoch-based sleep scoring, several methodological limitations warrant consideration. Firstly, this study focused on sleep and wake-like classification rather than multi-class sleep staging, a choice informed by the availability constraints of temporally continuous manual scoring across all sleep stages. Feedback from the expert scorer who conducted temporally continuous sleep-wake scoring in the Austin data, highlighted difficulties of such detailed manual annotation, particularly due to the lack of facilitative tools. The model is inherently adaptable to include multi-class high-frequency sleep staging; however, its expansion requires careful validation and depends on the availability of suitable data. Second, it is important to note that only one expert provided continuous scoring per dataset, which may introduce subjectivity into the manual reference labels. However, we note similar performance in the Harvard and Austin datasets, suggesting the model appears to have generalized well to data scored by different scorers. Evaluating the model on larger, heterogeneous datasets with similar temporally continuous scoring will further establish its scalability and effectiveness. Finally, participant-wise performance revealed two outliers in the Austin dataset (Figure 3). We investigated whether these participants exhibited any notably different demographic or clinical characteristics compared to the rest, but found no clear differences. However, we noted that the same participants showed the largest discrepancies in TST between conventional AASM-based manual scoring and temporally continuous manual scoring.

### 4.4 Conclusions

This study presents a deep learning-based high-temporal-resolution sleep-wake classification model that consolidates wakefulness and arousals. By using temporally continuous manual scoring for validation, the study addresses key limitations of the conventional 30-second epoch-based sleep scoring practice and marks an important advance toward more physiologically consistent sleep assessment. The model demonstrated high concordance with the temporally continuous manual annotations and showed strong correlation for key sleep-wake metrics, such as TST and sleep-to-wake transitions. Together, these results lay a strong foundation for physiologically meaningful sleep analysis with potential diagnostic and prognostic relevance.

#### Data availability

The MESA Sleep dataset used in this study is publicly available through the National Sleep Research Resource (NSRR) at https://sleepdata.org/datasets/mesa. Access is provided under the NSRR data use agreements. The Harvard and Austin datasets analyzed in this study are not publicly available due to institutional restrictions. Researchers interested in accessing these datasets may contact the corresponding author with a reasonable request, which will be considered in line with institutional approvals and data-sharing policies.

#### Financial disclosure

HK was supported by the State Research Funding for university-level health research, Kuopio University Hospital, Wellbeing Service County of North Savo (5041803). TL received research funding from the Research Council of Finland (361199) and from the State Research Funding for university-level health research, Kuopio University Hospital, Wellbeing Service County of North Savo (5041820). JT received research funding from the Sigrid Juselius Foundation and has received royalties related to the Bittium BrainStatus EEG electrode set (patents EP2916730A1, WO2014072582A1, US9770184B2). SS was funded by the NIH NHLBI (R01HL168067) and received grant support from Apnimed, Inspire Medical Systems, Prosomnus, Dynaflex, and SleepRes; served as a consultant for Incannex (Advisory Board), Apnimed, Nox Medical, Eli Lilly, and Mosanna; is co-inventor of intellectual property related to combination pharmacotherapy for sleep apnea (patented, licensed, royalties) and wearable sleep apnea phenotyping (patented); and holds equity in Achaemenid. DLM was supported by an NHMRC Ideas Grant (2001729) and an NHMRC EU-Horizons Grant (2007001). PIT was supported by the Australian NHMRC (2001729 and 2007001).

All other authors report no funding and no financial competing interests.

#### Non-financial disclosure

The authors declare no conflicts of interest.

