## Supplementary material for "Temporally Continuous Automated Sleep-Wake Classification Using Deep Learning": Online Supplement Material

### Details of Neural Network Architecture

The architecture used in this study is a modified version of the publicly available U-Sleep algorithm [1], a fully convolutional, feed-forward deep neural network designed for sleep staging. U-Sleep is based on the widely used U-Net framework [2]. Originally proposed for biomedical image segmentation, U-Net typically features a symmetric encoder-decoder architecture. The encoder progressively reduces spatial or temporal resolution to extract high-level features, while the decoder reconstructs the output by gradually restoring resolution [2]. A central feature of U-Net is the use of skip connections between corresponding encoder and decoder layers, enabling the integration of coarse semantic features with fine-grained details from earlier layers. These skip connections help preserve important information that may otherwise be lost during down-sampling and contribute to more accurate, temporally aligned outputs [2]. Although initially developed for two-dimensional data, U-Net has since been adapted for one-dimensional inputs in various time-series applications [3,4]. One of its main strengths is its ability to generate high-resolution outputs, allowing precise spatial localization in imaging and fine-grained temporal predictions in time-series data [2,4]. Given U-Sleep's foundation in a U-Net variant inherently suited for high-temporal-resolution predictions, along with its demonstrated generalizability across multiple datasets, we adopted the U-Sleep architecture with targeted modifications to better meet the objectives of our study.

The final network consisted of three main modules: encoder, decoder, and segment classifier. The encoder module extracted abstract features from the input signals while reducing their temporal resolution. To account for the comparatively smaller data size used in the present analysis relative to the original U-Sleep study, the number of encoder blocks was reduced from 12 to 8. Each encoder block consisted of a one-dimensional convolution layer with a kernel size of 7, reduced from the original size of 9, to simplify the architecture and minimize the risk of overfitting associated with the relatively smaller training data. Kernel dilation was not used, and the stride was set to 1 in the convolutional layer, consistent with the original implementation. Each block also included an Exponential Linear Unit (ELU) activation function. As in the original U-Sleep model, batch normalization was applied after each convolutional layer to improve convergence stability, and max-pooling with a kernel size and stride of 2 was used to progressively down-sample the feature maps. A reduced complexity factor of 1.5 (instead of 1.67) was also applied. These adjustments further contributed to improving computational efficiency while preserving the model's ability to extract meaningful features.

The decoder module reconstructed the high-level feature representations back to the original temporal resolution while retaining the learned hierarchical features. To maintain consistency with the encoder and reduce model complexity in accordance with the relatively smaller training data, the decoder was also designed to comprise 8 blocks, reduced from 12 in the original U-Sleep implementation. Each decoder block performed nearest-neighbor up-sampling with a kernel size of 2, followed by a convolution operation with a kernel size of 2 and a stride of 1, consistent with the original design. Subsequently, the ELU activation function and batch normalization were also retained. The up-sampled features were concatenated with the batch-normalized feature maps (prior to max pooling) from the corresponding encoder block at matching temporal resolutions (e.g., first decoder block matching the last encoder block). A one-dimensional convolution layer, non-linearity, and batch normalization were then applied to refine the combined features, resulting in outputs at 128 Hz that represented latent sleep-wake states.

The segment classifier module processed the intermediate high-frequency decoder outputs to generate sleep-wake predictions at a 1-second resolution, which could be adjusted if needed. It applied average pooling per channel over defined segment windows, followed by two pointwise convolution layers with a kernel width and stride of 1, with ELU activation in the first layer. This configuration enabled the model to learn a non-linear weighted combination of the average scores across the interval. Finally, a softmax layer converted the scores into probabilistic sleep-wake predictions. The full network was trained in an end-to-end manner.

Additional technical modifications were implemented to accommodate differences in input data formats and to enable model training using shorter epoch durations. For example, while the original U-Sleep implementation supported multiple input formats for initial data processing, including extensible markup language (XML)-based sleep stage annotations, the XML schema used in one of our datasets differed from that of the original pipeline and required structural adaptation. Similarly, the Harvard dataset we had access to, included PSG recordings only in a custom MATLAB data format (these PSGs were originally recorded using the Spike software but were then processed into a Matlab-based format), necessitating further modifications to integrate these data files into the study workflow. Additional adjustments were also made to ensure compatibility with the coding environment used in this study. Most other settings and hyperparameters were retained from the original U-Sleep study for consistency.

For the initial model training with the Multi-Ethnic Study of Atherosclerosis (MESA) dataset, we used a learning rate of $1 \times 10^{-6}$ and a batch size of 64, optimizing the model with the Adam optimizer. Early stopping was implemented to halt training if validation performance plateaued for 25 consecutive training cycles, and a ModelCheckpoint callback was used to save the best-performing model. No regularization was applied during this phase, as overfitting was negligible.

An overview of the neural network architecture is presented in Figure S1.

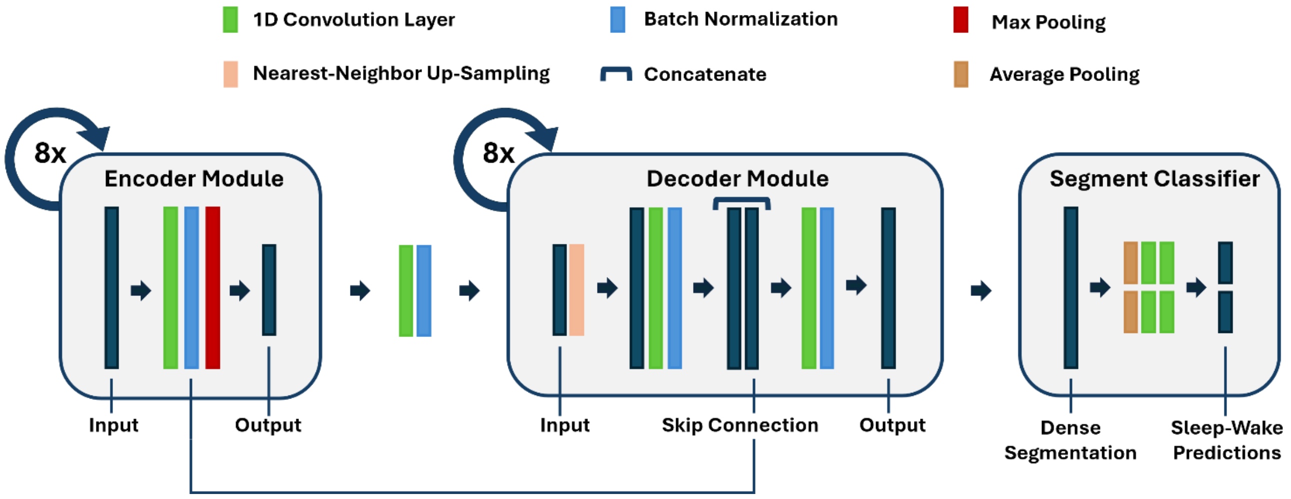


***Figure S1:*** *Overview of the neural network architecture. The network consists of three main modules: encoder, decoder, and segment classifier. 1D = One-dimensional.*

### Details of Transfer Learning Procedure

To adapt and further fine-tune the algorithm for temporally continuous scoring, transfer learning was employed. The MESA pre-trained 1-second model with arousal incorporation and heuristic correction steps served as the base model. During fine-tuning, all layers of the base model were frozen except for the last three, which were selectively trained to align with the features of the temporally continuous sleep–wake scoring. Input signals from the Harvard dataset comprised C3-A2, C4-A1, O2-A1, F3-A2, LOC-A2, and ROC-A1 channels, which included internal high-pass (0.3 Hz) and low-pass (35 Hz) filters and were resampled from 125 Hz to 128 Hz, similar to MESA. The Austin dataset input signals included F4-M1, C4-M1, O2-M1, E1-M2, and E2-M2 channels. These signals were also resampled from 256 Hz to 128 Hz and preprocessed using 4th-order Butterworth high-pass and low-pass filters at 0.3 Hz and 35 Hz, respectively, to ensure consistency across datasets.

The fine-tuning process incorporated regularization and optimization techniques to prevent overfitting. Dropout layers were applied with a rate of 0.5, and L2 regularization ($1 \times 10^{-3}$) was used for kernel weights. The initial learning rate was set to $1 \times 10^{-3}$. Early stopping was triggered if validation performance plateaued for 50 consecutive training cycles and the learning rate was adaptively reduced by a factor of 0.1 after 20 cycles of stagnant validation performance, with a minimum learning rate of $1 \times 10^{-7}$. Moreover, during training, Gaussian noise ($\mu$=0, $\sigma$=0.01) was added to simulate natural variations, and random temporal shifts were applied to input segments by up to 10% of their duration.

### Total Sleep Time (TST) and Sleep-to-Wake Transition Index Comparison After Probability-Based Heuristic Smoothing

Provided the minimum duration criteria used during manual arousal scoring in the Harvard dataset (>3 seconds with no upper limit), we implemented an additional post-hoc heuristic correction to address isolated 1-second sleep or wake predictions produced by the model. These isolated segments were defined as a single 1-second classification surrounded by the opposite class. For each such instance, we examined a 3-second window centered on the segment and computed the mean prediction probability for the class assigned to the 1-second segment. If this mean probability was below 0.5, the 1-second segment was smoothed to match the surrounding class. If the probability was at least 0.5, the full 3-second window was reassigned to the class of the central 1-second segment.

This correction had a modest but meaningful impact on the agreement metrics. The correlation between the model and the temporally continuous scoring for the sleep-to-wake transition index improved to 0.72 ($p<0.001$), and the mean difference in the Bland-Altman analysis was reduced to 0.19 (Figure S2). Other performance measures remained largely unchanged, indicating that this heuristic adjustment selectively further addressed some misaligning 1-second standalone segments without altering the broader pattern of results.

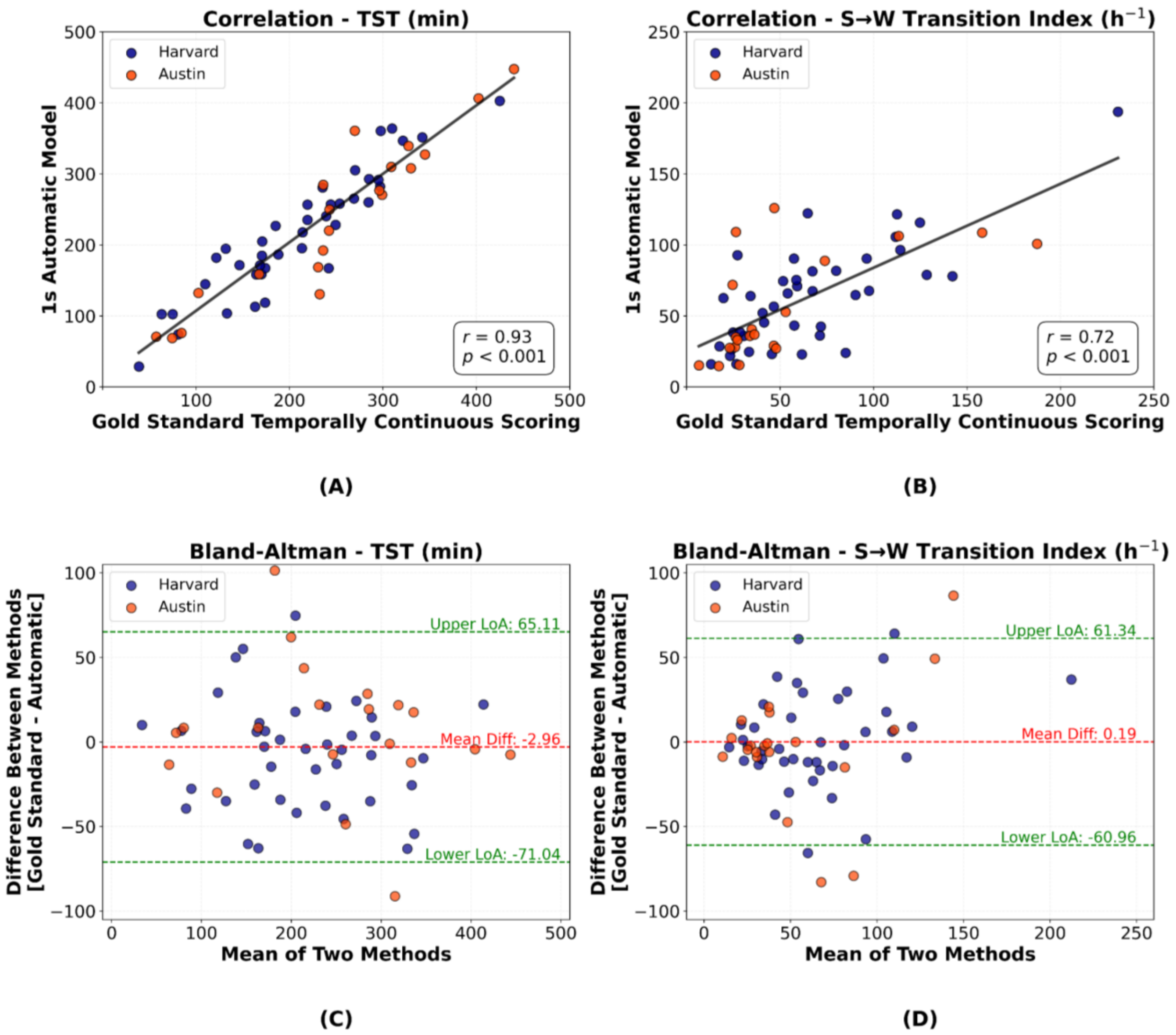


***Figure S2:*** *Analyses of TST and sleep-to-wake transition indices between the 1-second automatic method and the gold standard temporally continuous scoring after probability-based heuristic smoothing.* ***(A)*** *Correlation plot of TST (r=0.93, p<0.001).* ***(B)*** *Correlation plot of sleep-to-wake transition index (r=0.72, p<0.001).* ***(C)*** *Bland-Altman plot for TST comparison.* ***(D)*** *Bland-Altman plot for sleep-to-wake transition index. TST = total sleep time, S→W = sleep-to-wake.*
